# Treatments administered to the first 9,152 reported cases of COVID19: a systematic review

**DOI:** 10.1101/2020.05.07.20073981

**Authors:** David C. Fajgenbaum, Johnson S. Khor, Alek Gorzewski, Mark-Avery Tamakloe, Victoria Powers, Joseph J. Kakkis, Mileva Repasky, Anne Taylor, Alexander Beschloss, Laura Hernandez-Miyares, Beatrice Go, Vivek Nimgaonkar, Madison S. McCarthy, Casey J. Kim, Anna Wing, Michael A. Mayer, Ruth-Anne Langan Pai, Sarah Frankl, Megan Fisher, James Germi, Cornelia A. Keyser, Philip Angelides, Ashwin Amurthur, Joanna Jiang, Rozena Rasheed, Eric Rodriguez-Lopez, Erin Napier, Bruna Martins, Stephen Bambury, Karen Gunderson, Nick Goodyear, Duncan Mackay, Sheila K. Pierson

## Abstract

The emergence of SARS-CoV-2/2019 novel coronavirus (COVID19) has created a global pandemic with no approved treatments or vaccines. Many treatments have already been administered to COVID19 patients but have not been systematically evaluated. We performed a systematic literature review to identify all treatments reported to be administered to COVID19 patients and assess time to clinically meaningful response for treatments with sufficient data. We searched PubMed, BioRxiv, MedRxiv, and ChinaXiv for articles reporting treatments for COVID19 patients published between 12/1/2019–3/27/2020. Data were analyzed descriptively. Of the 2,706 articles identified, 155 studies met inclusion criteria, comprising 9,152 patients from 14 different countries. The cohort was 45.4% female and 98.3% hospitalized and mean (SD) age was 44.4 years (SD 21.0). The most frequently administered drug classes were antivirals, antibiotics, and corticosteroids, and of the 115 reported drugs, the most frequently administered was combination lopinavir/ritonavir, which was associated with a time to clinically-meaningful response (complete symptom resolution or hospital discharge) of 11.7 (1.09) days. There was insufficient data to compare across treatments. A large number of treatments have been administered to the first 9,152 reported cases of COVID19. These data serve as the basis for an open-source registry of all reported treatments given to COVID19 patients. Further work is needed to prioritize drugs for investigation in well-controlled clinical trials and treatment protocols.

## Introduction

SARS-CoV-2 and its related disease, 2019 novel coronavirus (COVID19), is a global pandemic. Over 2,200,000 cases and 150,000 deaths have been reported to date worldwide.^[1]^ With an R_0_ reportedly as high as 2-2.5 and a significant case fatality rate, this virus will continue to have a major impact on the health and well-being of humans worldwide.

COVID19 patients exhibit a highly heterogeneous clinical course from mild flu-like symptoms to cytokine-storm driven acute respiratory and multi-organ failure.^[2]^ Physicians worldwide have administered numerous treatments off-label and through clinical trials to treat COVID19 patients. Although only four months have elapsed since the emergence of COVID-19, many case reports, single center series, and interventional studies have been published in medical journals as well as pre-publication archives. Some of these treatments have received widespread attention and are currently undergoing randomized controlled trials while others have not. Identifying and inventorying the full range of treatments reported in use is critical for physicians treating COVID19. Further indicators of effectiveness among those treatments is important for governments, public health organizations, and pharmaceutical companies identifying and prioritizing treatments for well-controlled clinical trials.^[3]^ However, a systematic effort to consolidate and centralize all treatment data is missing.

We performed a systematic review to inventory all treatments that have been given to COVID19 patients and to investigate response when possible.

## Methods

### Search strategy and selection criteria

We completed our systematic literature review according to the PRISMA guidelines.^[4]^ We searched PubMed, BioRxiv, MedRxiv, and ChinaXiv from Dec 1, 2019 to March 27, 2020 using the following terms “COVID19” OR “SARS-CoV-2” OR “2019-nCoV”. The search was not restricted to publications in any language as translation tools were used for articles not written in English. However, 22 articles were either not able to be accessed, translated, or interpreted by the extractor. Additional references were identified through bibliography searches and review of articles written by study authors. Inclusion criteria included all studies reporting the use of any treatments in COVID19 patients. No studies were excluded based on participants, treatments, outcomes, study design, or length of study. Both patient-level and summary-level studies were included. Given the urgency and importance of this information, articles deposited into online pre-publication archives were also included. The full text of all articles was reviewed by at least one data extractor to determine if the paper met the inclusion and exclusion criteria. A review protocol can be accessed at https://www.med.upenn.edu/CSTL/drug-repurposing.html.

### Data extraction

All articles meeting inclusion criteria were read and extracted into a centralized spreadsheet. Elements collected included article type, nationality of the authors, patient disease characteristics, treatments administered, and outcomes, when available. A second individual, either holding a medical degree or currently in medical school, performed an independent review of every article and repeated data extraction for every data point. A third individual (JSK) resolved discrepancies between the two extractors. JSK also reviewed the article list and data extractions to remove duplicates and resolve discrepancies related to study inclusion. Data on all patients with drug treatment information available was included to avoid selective reporting bias within studies, but publication bias is likely present in this study.

### Data analysis

Data were analyzed descriptively; no hypothesis tests were performed. Frequency counts and percentages were used to describe categorical data. A weighted mean (standard deviation) was tabulated for continuous data (age, C-reactive protein, time to clinically meaningful response). For summary-level studies that presented continuous data using a median and interquartile range or sample range, sample mean and standard deviation were imputed using the Wan et al. method.^[5]^ Data were combined, and an inverse-variance weighted mean was calculated for age and time to clinically meaningful response. Time to clinically meaningful response (TCMR) was calculated when available at a per-patient and per-drug level and was defined as the shortest duration between drug start and full symptom resolution (according to author) or hospital discharge. When data on both of these time points were provided, the shortest duration was considered as the TCMR. Treatments were classified according to the Anatomical Therapeutic Classification (ATC) therapeutic subgroup. Analyses were performed using R (version 3.6.0).

## Results

We identified and reviewed 2,706 papers (PubMed: 1795, Biorxiv: 298, Medrxiv: 591, Chinaxiv: 22). Of those, 2,542 were excluded and 164 underwent complete data extraction by two independent extractors (Figure 1). Reasons for exclusion include duplication and lack of information on drug treatments administered to COVID19 patients. The studies varied in category from single case reports in pre-publication archives to published randomized controlled trials. 155 studies from 14 countries (Afghanistan, China, France, Germany, Italy, Japan, Korea, Scotland, Singapore, Spain, Taiwan, United Kingdom, United States, and Vietnam) satisfied inclusion criteria; 9 articles were determined not to meet inclusion criteria when re-reviewed by a second extractor. Of the 155 studies, 117 were published in journals indexed on PubMed; the remainder were published in online archives. Studies reviewed included 87 single-patient retrospective case reports, 66 retrospective case series of between 2 and 1,099 cases, and 2 interventional clinical trials. Data for 9,152 patients were reported, with data reported from 238 patients on an individual patient-level and 8,914 patients on a summary level. These data are available in Supplementary Table 1 and a registry of COVID19 treatments can be accessed at https://www.med.upenn.edu/CSTL/drug-repurposing.html.

**Figure 1.**
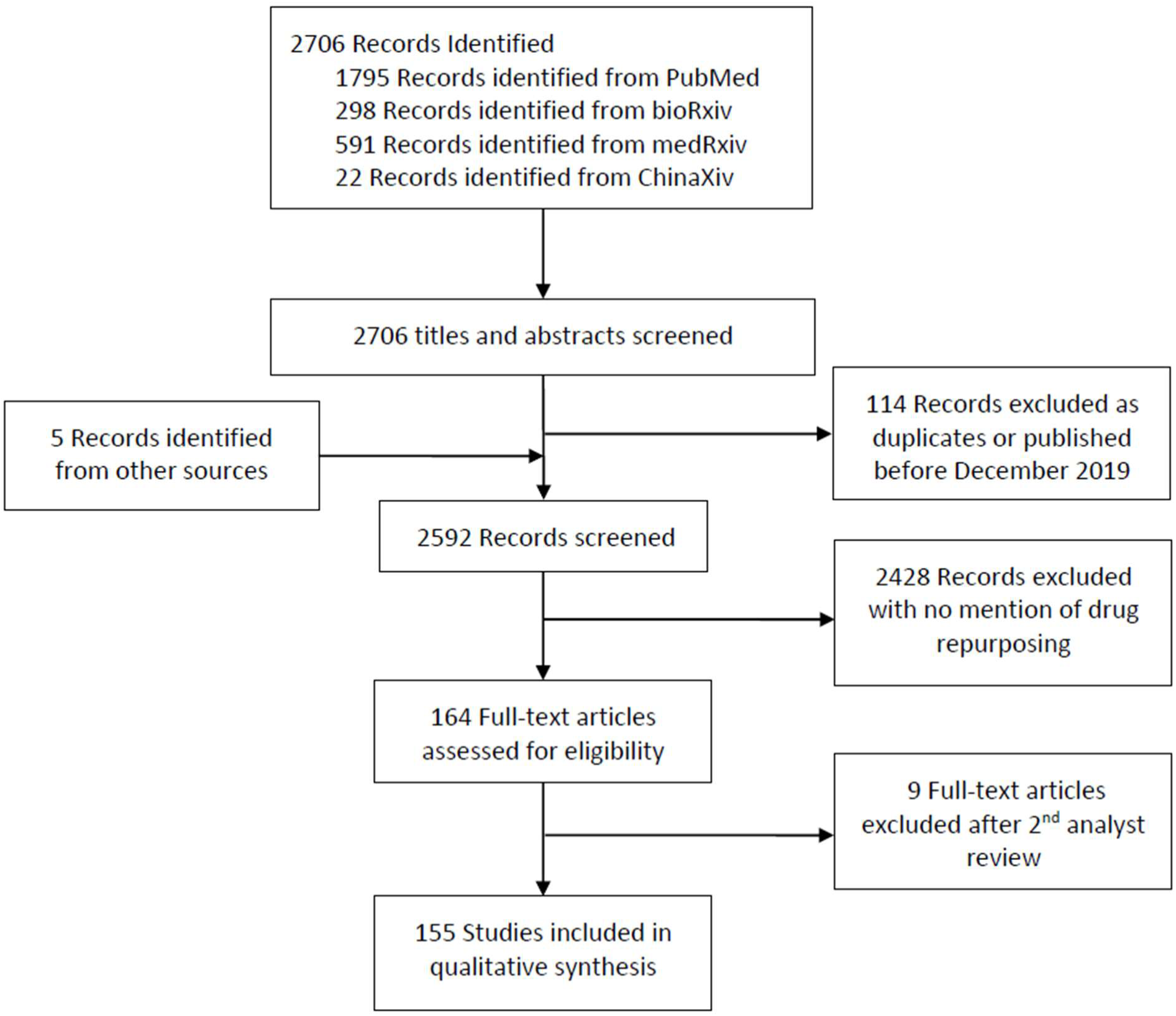
PRISMA study selection.

Patients from at least 14 different countries were represented (Table 1). The weighted mean patient age was 44.4 (SD 21.0), and 45.4% of patients were female. The method of positive COVID19 testing was reported for 91.3% of patients included in the analysis. Nearly all cases were reported to be hospitalized (98.3%), and 1672 (18.3%) were reported to require ventilation. In 2,470 (27.0%) cases, the author described the patient(s) as having “severe” disease.

**Table 1.**
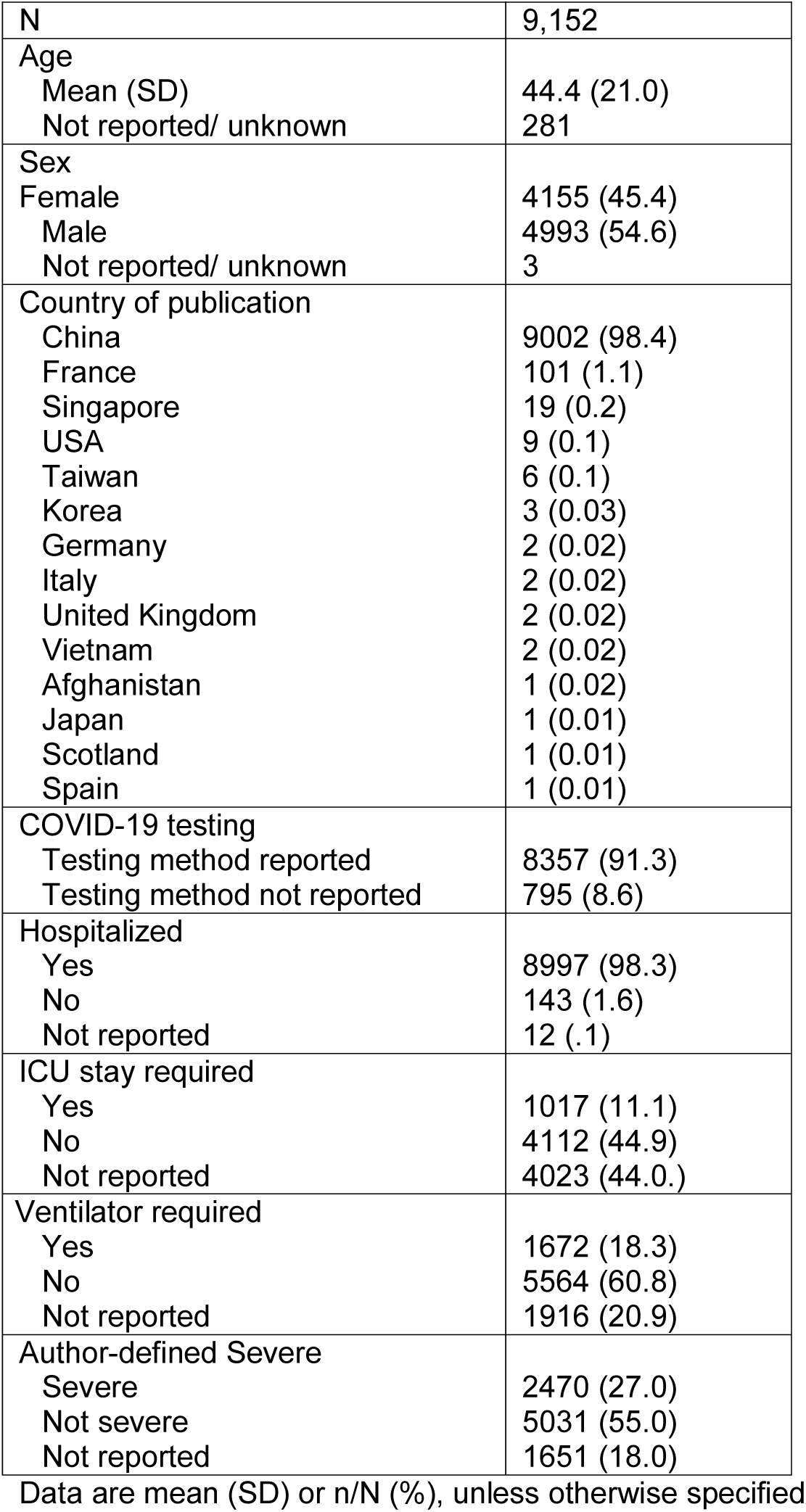
Study populations

|  |  |
| --- | --- |
| N | 9,152 |
| Age |  |
| Mean (SD) | 44.4 (21.0) |
| Not reported/ unknown | 281 |
| Sex |  |
| Female | 4155 (45.4) |
| Male | 4993 (54.6) |
| Not reported/ unknown | 3 |
| Country of publication |  |
| China | 9002 (98.4) |
| France | 101 (1.1) |
| Singapore | 19 (0.2) |
| USA | 9 (0.1) |
| Taiwan | 6 (0.1) |
| Korea | 3 (0.03) |
| Germany | 2 (0.02) |
| Italy | 2 (0.02) |
| United Kingdom | 2 (0.02) |
| Vietnam | 2 (0.02) |
| Afghanistan | 1 (0.02) |
| Japan | 1 (0.01) |
| Scotland | 1 (0.01) |
| Spain | 1 (0.01) |
| COVID-19 testing |  |
| Testing method reported | 8357 (91.3) |
| Testing method not reported | 795 (8.6) |
| Hospitalized |  |
| Yes | 8997 (98.3) |
| No | 143 (1.6) |
| Not reported | 12 (.1) |
| ICU stay required |  |
| Yes | 1017 (11.1) |
| No | 4112 (44.9) |
| Not reported | 4023 (44.0.) |
| Ventilator required |  |
| Yes | 1672 (18.3) |
| No | 5564 (60.8) |
| Not reported | 1916 (20.9) |
| Author-defined Severe |  |
| Severe | 2470 (27.0) |
| Not severe | 5031 (55.0) |
| Not reported | 1651 (18.0) |

Data are mean (SD) or n/N (%), unless otherwise specified

All patients included in this analysis received at least one treatment intended to treat COVID19 (Table 1). Fourteen therapeutic categories comprised a total of 115 reported treatments, as well as many non-descript treatments (e.g. antibiotics not otherwise specified”). Treatments described were administered alone, concurrently, or sequentially with others. Given the nature of the reports, we did not differentiate concurrent or sequential treatment regimens. The most frequently administered classification of treatments were antivirals (N=6547, 71.5%), antibiotics (N=4263, 46.6%), and corticosteroids (N=2392, 26.1%) (Figure 2A). The most frequently administered treatment given to all patients was combination lopinavir/ritonavir (N=2000, 21.9%), followed by interferon α/β (N=1767, 19.3%) and immunoglobulins (N=1049, 11.5%). (Figure 2B). Of the treatments identified, 100 (86.9%) were administered to fewer than 1% of all patients.

**Figure 2.**
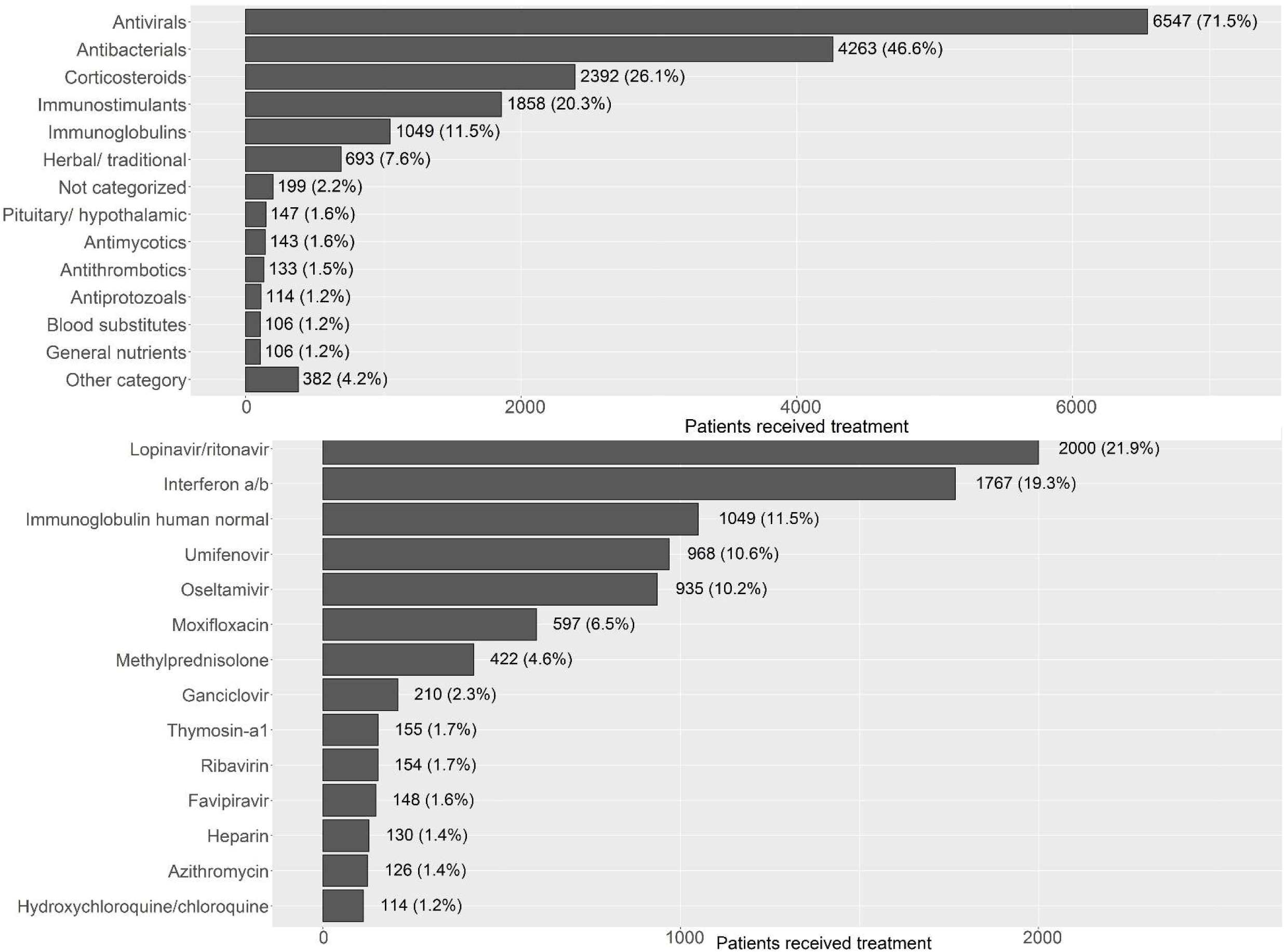
Most frequently administered therapeutic classes of drugs. Most frequently administered therapeutic classes of drugs according to the Anatomical Therapeutic Classification system (A), and most frequently administered individual drugs reportedly used among the 9,152 patients (B).

On a per drug basis, response data were sparse and were largely only available on the individual patient-level (e.g. case study). When available, we calculated duration between initiation of treatment and clinically-meaningful response (time to clinically-meaningful response, TCMR). Of note, drugs were included in the calculation regardless of whether they were used alone, concurrently, or sequentially, and therefore response time cannot be conclusively attributed to a given drug. On an individual treatment-level, only 6 treatments included patient TCMR data for at least 10 patients and from at least 10 studies. Interferon-α/β, which had the highest amount of available response data (N=107 patients from N=14 studies), was associated with the shortest weighted TCMR, at mean (SD) 9.9 (2.65). Combination lopinavir/ritonavir included the second most available data (N=76 patients from N=15 studies) and was associated with a TCMR of 11.7 (1.09) days. From our review, oseltamivir (10 patients from 10 studies) was associated with the longest average TCMR of 19.8 (10.62), though the sample with available data was limited (Table 2, Figure 3). We did not compare drugs as sample sizes are small, and well-controlled clinical trials are needed to determine efficacy of these drugs.

**Table 2.**
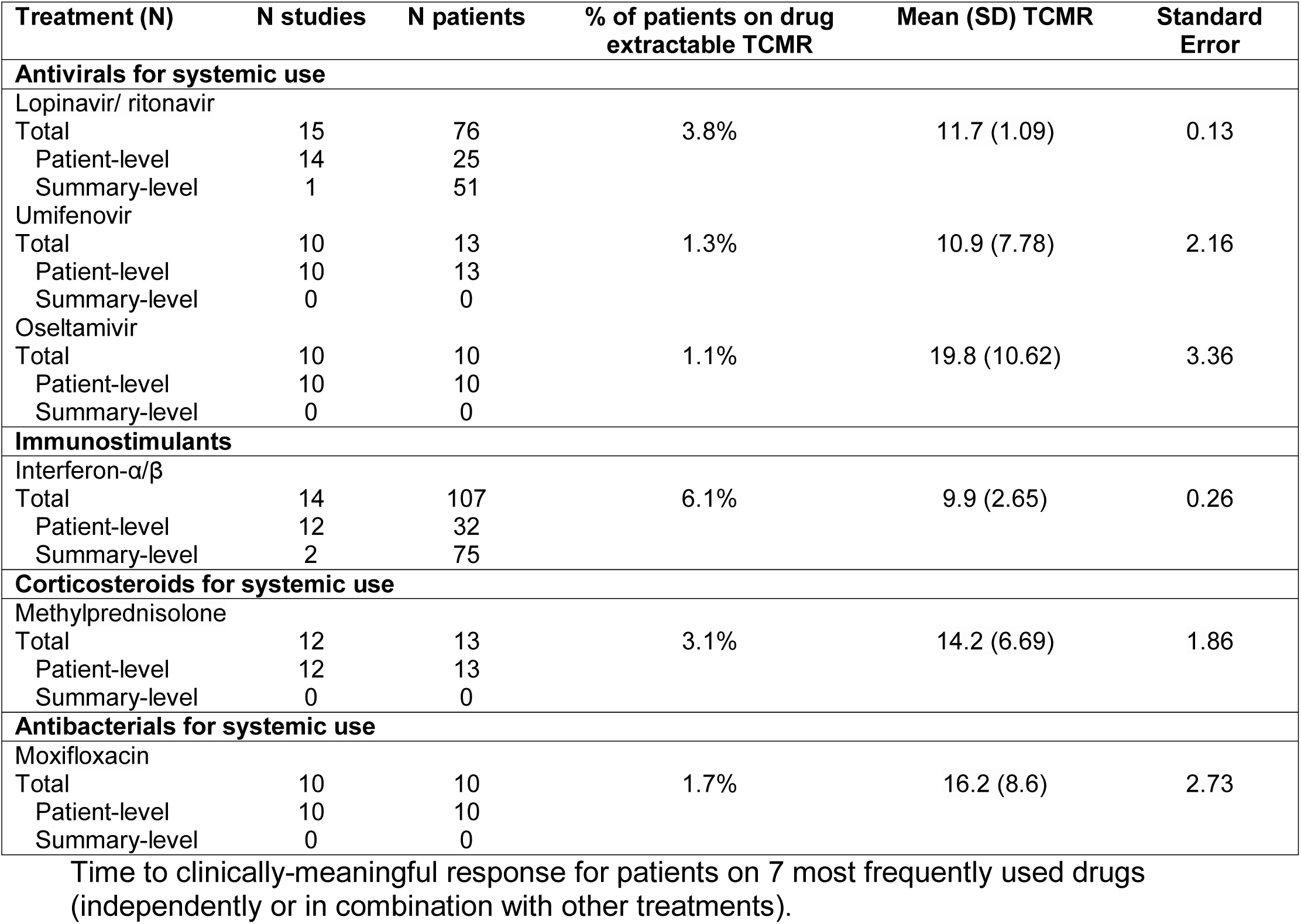
Time to clinically-meaningful response for patients on 7 most frequently used drugs (independently or in combination with other treatments).

**Figure 3.**
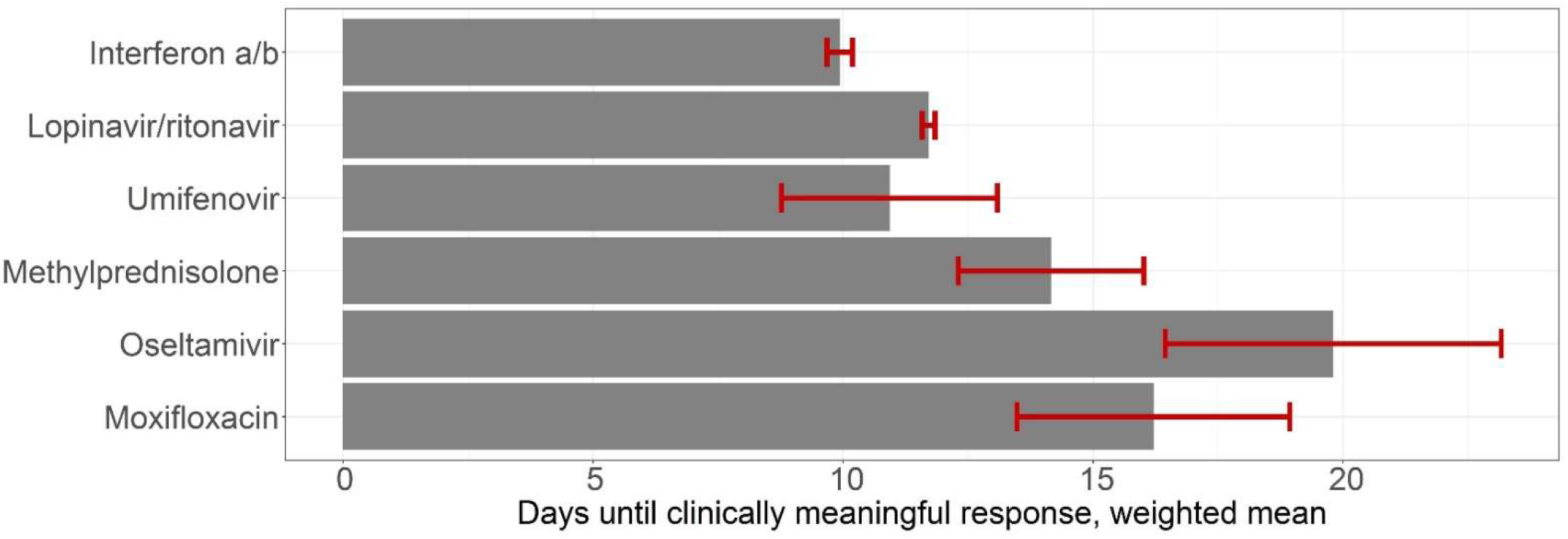
Mean weighted time to clinically meaningful response associated with the most frequently used drugs. Clinically meaningful response was calculated as the duration from drug start until the earliest of author-reported resolution of symptoms or hospital discharge. Data on drugs with response times available from at least 10 studies are included. A mean and standard deviation was imputed from summary-level reports with median (interquartile range), and a weighted mean was calculated from available data. Drugs may have been used along or concurrently with others. TCMR observations were available for <15 patients for oseltamivir, methylprednisolone, moxifloxacin, and umifenovir.

For the two most frequently used drugs, we restricted to the studies reporting patient-level observations and plotted a Kaplan-Meier curve for time to clinically meaningful response (N=37 observations for interferon-α/β, and N=34 observations for lopinavir/ritonavir). Patients were included if TCMR or author-reported time to improvement was available. Patients who were not reported to achieve our definition of clinically-meaningful response were censored at time to improvement (Supplementary Figure 1). For both drugs, median TCMR was less than 2 weeks.

Recent insights into COVID19 pathogenesis suggest several potential mechanisms of action for the treatments identified in this study (Figure 4). SARS-CoV-2 binds to the ACE2 receptor on ciliated bronchial epithelial cells to gain entry into these cells for viral replication and dissemination in the airway and systemically.^[6]^ Patients with a weakened immune response would be expected to have difficulty controlling COVID19. Patients with a hyper-immune response experience acute respiratory distress syndrome, septic shock, and multiple organ system failure due to a cytokine storm.^[7]^ Treatments used to date have been proposed to work by: 1) limiting entry into ciliated bronchial epithelial cells (N-acetylcysteine, heparin, meplazumab, umifenovir, hydroxychloroquine), 2) inhibiting viral replication (interferon-α/β, ritonavir/lopinavir, oseltamivir, ganciclovir, ribavirin, favipiravir, remdesivir, danoprevir), 3) preventing viral dissemination via antibody-mediated neutralization by increasing SARS-CoV-2-specific antibodies (convalescent plasma) or non-specific antibodies (IVIg, thymopentin), 4) strengthening a weakened immune response with immunostimulants (interferon-α/β, thymosin-α-1), 5) preventing a hyper-immune response with immunosuppressants (corticosteroids, hydroxychloroquine, IVIg), or 6) controlling a hyper-immune response (corticosteroids, tocilizumab).^[8–11]^ A number of antibiotics were also given to COVID19 patients. These drugs may be acting by preventing secondary infections, controlling inflammation, modulating the microbiome, or directly having an anti-viral effect; they may also have had no effect.

**Figure 4.**
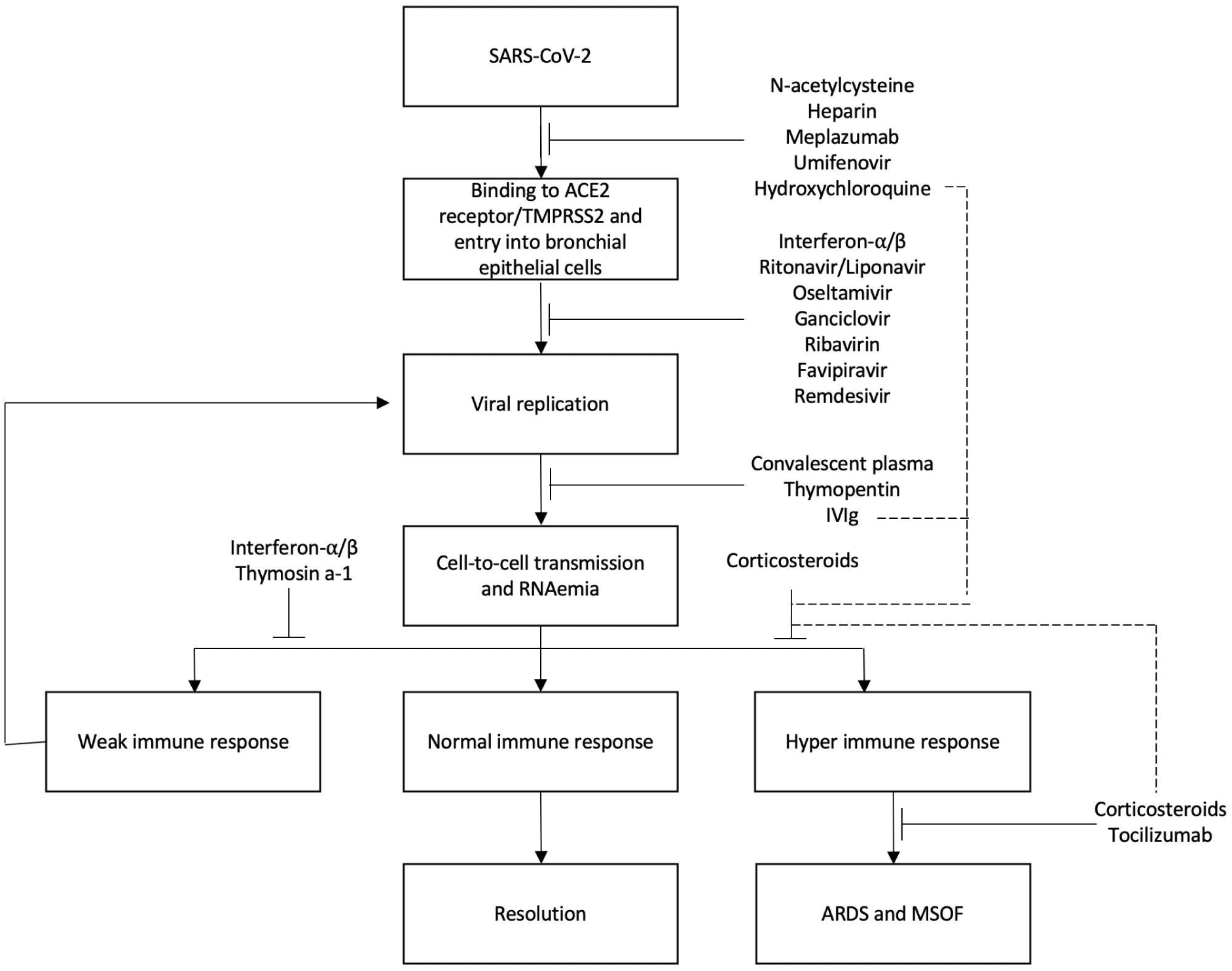
Potential mechanisms of treatments used to date against COVID19. Antibiotics such as azithromycin, moxifloxacin, ceftriaxone, and cefoperazone were used more than 25 times but the potential mechanism of action is unknown. A number of treatments were also administered in this study support vital organ function, such as bacillis lichenformis (gastrointestinal), antacids (gastrointestinal), continuous renal replacement therapy (kidneys), vasopressors/vasocontrictive agents (cardiovascular), and expectorant and antitussive agents (respiratory). IVIg, intravenous immunoglobulin. Dotted lines represent potential secondary mechanisms of action.

The treatments that have received the most attention to date include hydroxychloroquine, azithromycin, antivirals used effectively against similar viruses (SARS, MERS, influenza), convalescent plasma, and cytokine storm-directed therapies. In our systematic review, hydroxychloroquine was used 114 times. These patients are from a small case series and two large observational studies, which did not provide enough information to calculate a TCMR. Hydroxychloroquine is believed to increase endosomal pH needed for virus-cell fusion, interfere with glycosylation of the ACE2 receptor, and modulate the immune response.^[12]^ Ritonavir/lopinavir, administered 2000 times in this study, is a protease inhibitor approved for HIV that has demonstrated activity against MERS and SARS.^[13,14]^ Despite its frequent use, one randomized controlled trial included in this study reported no benefit beyond standard of care. However, the study began administration more than 10 days into the disease course, so it is still being studied earlier in disease course.^[15]^ Interferon, the second-most frequently administered agent in this study, is a key anti-viral cytokine produced by the host immune system that can inhibit coronavirus replication and boost the immune response.^[16–18]^ It was shown to be effective against SARS-CoV and MERS-CoV.^[19,20]^ Oseltamivir was also used frequently in this study, though the primary intent was likely to prevent concomitant influenza infection and it does not seem to be a promising treatment approach. Remdesivir, a nucleoside inhibitor not yet approved for any indication has demonstrated positive *in vitro* activity; it was used in a small number of cases.^[12]^ Umifenovir, approved for influenza in China and Russia, demonstrated a TCMR of 10.9 (7.78) days in this study, though sample size was low. A study of 67 patients revealed decreased mortality and improved discharge in patients treated with umifenovir.^[21]^ Interestingly, a randomized controlled trial demonstrated greater clinical improvement in favipiravir-treated patients than umifenovir among moderately ill patients.^[22]^ Convalescent plasma was only given in 12 cases, but it was used previously to treat MERS and SARS and a number of studies are underway.^[23–25]^

## Discussion

Despite advances in medical care, therapeutics, and infrastructure that have lowered the burden of infectious diseases in recent years, COVID19 has emerged as a leading cause of death in developed and developing countries. Drug repurposing is the fastest route towards an effective and accessible treatment against COVID19 before a vaccine is available. A previously unquantified but large number of treatments have been tried off-label or experimentally. To date, only small case reports and single-center studies have reported off-label treatments and data on their potential effectiveness. Some of these publications have received more attention than others leading to further off-label use. It is important to systematically evaluate all previously used treatments to avoid missing effective options. In this systematic review, we identified 115 reported treatments that have been used off-label or experimentally to treat COVID19; we report an initial assessment of associations with clinically meaningful response. Unsurprisingly, antivirals were the most frequently administered class of treatments. Combination lopinavir/ritonavir and interferon-α/β were the most frequent treatments given to all patients. Given the small amount of data and the fact that drugs are often given concurrently or sequentially, we did not seek to compare drugs, but lopinavir/ritonavir and interferon-α/β, which had the most amount of data, were each associated with average TCMR of less than 2 weeks.

These data can be used to prioritize promising treatments for randomized controlled trials. Given that the natural history of COVID19 is complete resolution in most patients, it is essential that prospective, randomly assigned control groups are used to compare with interventional groups. Further, this study can inform public health organizations, governments, and treating physicians about treatments that have been used and could be considered in future patients, considering the current absence of randomized controlled trial data. Many of the 76 regimens proposed by the World Health Organization for COVID19 treatment in February 2020, as well as proposed in Chinese governmental guidelines, include treatments found in this study.^[3, 5]^ These drugs were likely often given because they were included in these guidelines. Also, the current case fatality rate of COVID19 is only interpretable in the context of the medical care and treatments provided to patients to date. Some of the most frequently administered treatments in this study could potentially serve as a starting point for a list of essential medicines for resource-limited regions. Lastly, there are a number of high throughput drug screening efforts underway to identify existing drugs that may have activity against SARS-CoV-2. These data provide information on drugs currently in frequent use.

Experience from treating similar cytokine storm disorders, such as idiopathic multicentric Castleman disease, hemophagocytic lymphohistiocytosis, and chimeric antigen receptor therapy, by targeting interleukin-6 (IL-6), IL-1, JAK/STAT, NFkB, mTOR, and NFAT suggest that drugs like siltuximab, anakinra, and tocilizumab may be effective for controlling the COVID19-related cytokine storm.^[7]^ The potential benefits for these drugs to control or prevent a hyper-immune response must be weighed against the risk of accelerating disease progression by suppressing the immune response. There are also unanticipated effects when treatments are deployed against novel diseases, such as the high incidence of venous thromboembolism with IVIg during the 2003 SARS epidemic.^[24–26]^

This study provides several other broad insights into COVID19. We observed 55%-to-45% male-to-female ratio in these moderate-to-severe COVID19 cases. A similar pattern was observed for SARS-CoV and MERS-CoV and suspected to be related to protection from sex hormones or differences in the burden of chronic diseases^. [29–36]^ The average age of 44.4 years is younger than many reports. This may be due to younger patients being more likely to survive a severe presentation and thus more likely to be written up as a case report. Pharmaceutical and traditional Chinese treatments were reported in these publications.^[37,38]^ We found that drugs classified as herbal and traditional medicine were used in 693 patients (7.6%). Public health organizations have recommended against corticosteroids for COVID19 unless indicated for other conditions, but they were some of the most frequently used treatments in our study.^[3,39,40]^ Further study is needed.

This systematic review has several important limitations. In view of the limited number of randomized controlled trials, all papers published in PubMed or archives were included. From our standpoint, data archived articles about patient characteristics and treatments were important and unlikely to change during the peer-review process. Given the current crisis, we chose to enlist a large number of extractors to review the 2,706 papers. To improve data quality, a medical doctor or medical student re-reviewed every single extracted case and any discrepancies were resolved by consensus among extractor #1, extractor #2, independent reviewer (JSK), senior statistician (SKP), and principal investigator (DCF). This sample set is highly skewed towards hospitalized patients and thus is not likely representative of all COVID19 patients, and publication bias is likely. Specifically, physicians tend to write up cases where patients have positive outcomes. Also, physicians with resources to write articles may have more resources available to treat their patients. Mortality was not assessed as the majority of publications did not report survival and many patients were still hospitalized at the time of publication. Instead, we used TCMR. This aggregated outcome helped us to overcome the heterogeneity of outcomes reported across studies. However, we did not consider the concurrent or sequential drug use when associating time to clinically meaningful response. Further, we could not control for the natural history or severity of the disease. These data mostly come from treatment of patients in Asia, and treatment strategies may differ in other regions. Finally, some studies were extracted through translations instead of the full original text. Despite these limitations, this systematic review provides the first broad overview of treatments tried against COVID19 and insights that can inform practicing public health organizations, clinicians, and clinical trial prioritization. Importantly, these data serve as the basis for an updated registry of all treatments reported to be given to COVID19 patients in the published literature (COvid Registry of Off-label & New Agents, CORONA; https://www.med.upenn.edu/CSTL/drug-repurposing.html).

COVID19 represents the largest global pandemic and widespread threat to human health in decades. Fortunately, thousands of drugs are available for other indications and can be used off-label immediately. Unfortunately, no central register of all treatments used for COVID19 exists, so limited data exist on treatment use and activity. Therefore, physicians do not have information on potentially promising treatments and biopharmaceutical companies must make decisions about clinical trials with limited data. We present the first systematic review of treatments used against COVID19. The importance of these data, heterogeneity of the data, and the time and resources required to perform this study emphasize the urgent need for systematic data collection on off-label and experimental treatments against COVID19 as well as randomized controlled trials of promising agents. Learning from this experience to inform future pandemics and aid in the discovery of effective treatments for the approximately 7,000 diseases without treatments is critically important.

## Data Availability

Data will be available at https://www.med.upenn.edu/CSTL/drug-repurposing.html.

## Acknowledgements

There was no funding source for this study. We wish to thank the physicians, researchers, and patients who contributed their data so as to help future patients.

## Contributors

JJK, MR, AT, AB, LHM, BG, VN, MSM, CJK, RAP, SF, PKA, JJ, RR, EN, DM reviewed and extracted data from articles and reviewed and revised the manuscript.

AG, MAT, VP reviewed and extracted data from articles, performed a re-review of every previously extracted articles, and reviewed and revised the manuscript

JSK did the literature search, reviewed and extracted data from articles, performed data quality queries, analyzed data, and reviewed and revised the manuscript.

SKP planned the study, analyzed data, and reviewed and revised the manuscript.

DCF planned the study and wrote the manuscript.

